# Exosomal microRNAs Drive Thrombosis in COVID-19

**DOI:** 10.1101/2020.06.16.20133256

**Authors:** Jessica Gambardella, Celestino Sardu, Marco Bruno Morelli, Vincenzo Messina, Vanessa Castellanos, Raffaele Marfella, Paolo Maggi, Giuseppe Paolisso, Xujun Wang, Gaetano Santulli

**Affiliations:** Department of Medicine, Einstein College of Medicine – Montefiore University Hospital, New York, NY; Department of Advanced Biomedical Sciences, International Translational Research and Medical Education (ITME), “Federico II” University, Naples, Italy; Department of Advanced Medical and Surgical Sciences, University of Campania, Naples, Italy; International University of Health and Medical Sciences “Saint Camillus”, Rome, Italy; Department of Infectious Diseases, University of Campania, Naples, Italy; Department of Molecular Pharmacology, Montefiore University Hospital, New York, NY

## Abstract

Thrombotic and thromboembolic complications have been shown to play a critical role in the clinical outcome of COVID-19. Emerging evidence has shown that exosomal miRNAs are functionally involved in a number of physiologic and pathologic processes. However, neither exosomes nor miRNAs have been hitherto investigated in COVID-19. To test the hypothesis that exosomal miRNAs are a key determinant of thrombosis in COVID-19, we enrolled patients positive for COVID-19. Circulating exosomes were isolated from equal amounts of serum and levels of exosomal miRNAs were quantified. We divided our population in two groups based on the serum level of D-dimer on admission. Strikingly, we found that exosomal miR-424 was significantly upregulated whereas exosomal miR-103a, miR-145, and miR-885 were significantly downregulated in patients in the high D-dimer group compared to patients in the low D-Dimer group (p<0.0001).

## Introduction

Coronavirus disease (COVID-19) is a public health crisis of global proportions caused by the *severe acute respiratory syndrome coronavirus 2* (SARS-CoV-2). Thrombotic and thromboembolic complications have been shown to play a critical role in the clinical outcome of COVID-19 (1-4).

MicroRNAs (miRNAs) are small non-coding RNAs that enhance mRNA degradation and/or inhibit protein translation (5,6). Emerging evidence has shown that exosomal miRNAs are functionally involved in a number of physiologic and pathologic processes (5,7,8).

However, neither exosomes nor miRNAs have been hitherto investigated in COVID-19.

## Methods

To test the hypothesis that exosomal miRNAs are a key determinant of thrombosis in COVID-19, we enrolled 26 patients positive for COVID-19 admitted to the Infectious Disease Departments of University of Naples “Vanvitelli” and San Sebastiano Caserta Hospital (Italy). The study was conducted according to the Declaration of Helsinki principles and approved by the local Ethical Committee. Written informed consent was obtained from all participants.

Circulating exosomes were isolated from equal amounts of serum, as previously described and validated by our group (9,10) and levels of exosomal miRNAs were quantified by RT-qPCR, normalizing values to spiked cel-miR-39 (10).

We divided our population in two groups based on the serum level of D-dimer on admission, using a previously published cut-off of 3 μg/ml (4).

## Results and Discussion

No significant differences in the main clinical characteristics were noted comparing patients with low (n=11, 72.7% male) *vs* high (n=15, 53.3% male) D-Dimer: age (59.7±22.7 *vs* 68.6±10.7), body mass index (27.3±2.3 *vs* 26.56±2.1 kg/m^2^), and hypertension (63.3% *vs* 66.7%).

Strikingly, we found a that exosomal miR-424 was significantly upregulated whereas exosomal miR-103a, miR-145, and miR-885 were significantly downregulated (**Figure 1, A-D**) in patients in the high D-dimer group compared to patients in the low D-Dimer group (*p<0*.*0001*). Regression analysis confirmed these findings (**Figure 1, E-H**).

**Figure 1.**
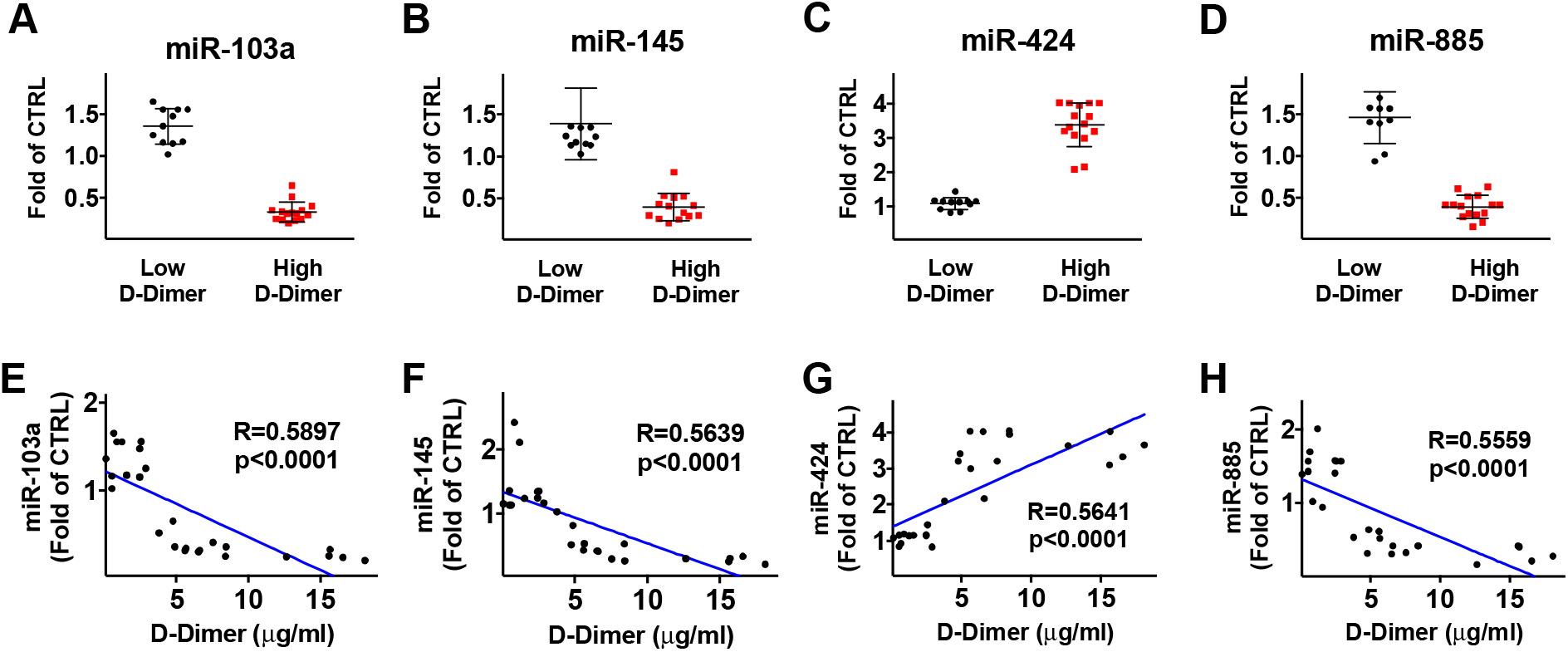
Expression levels of exosomal miRNAs in COVID-19 patients. Control (CTRL) indicates exogenous spiked-in cel-miR-39. In panels A-D, data are means±SD; p<0.0001.

Mechanistically, Tissue Factor has been identified as a direct target of miR-145, while miR-885 has been shown to target the von Willebrand Factor (5,11). Equally important, miR-424 has been associated with hypercoagulability whereas low levels of miR-103a have been observed in deep vein thrombosis (5,12), although precise mechanisms have not been fully defined for these miRNAs.

To our knowledge, this is the first study showing a functional contribution of exosomal non-coding RNA in COVID19. Limitations of our study include the relatively small population and the fact that we did not determine the exact source of exosomes; nevertheless, since endothelial dysfunction has been shown to be a prominent feature of COVID-19 and to contribute to the pro-thrombotic and pro-inflammatory state of the vasculature (2), we speculate that a main source could be represented by endothelial cells, which express these miRNAs in normal conditions (5).

## Data Availability

All data are available in the manuscript

